## Supplementary materials for "Cell-type-specific Alzheimer’s disease polygenic risk scores are associated with distinct disease processes in Alzheimer’s disease"

| Cell type | N_SNP_ROSMAP (%) | N_SNP_A4 (%) | N_SNP_HRC (%) | N_SNP_PRSet (%) |
|---|---|---|---|---|
| Ex | 78555 (7.6) | 81405 (7.6) | 445321 (6.8) | 20283 (10.7) |
| In | 101104 (9.7) | 104006 (9.7) | 578298 (8.8) | 25334 (13.3) |
| Ast | 82828 (8.0) | 85708 (8.0) | 458050 (7.0) | 21729 (11.4) |
| Mic | 71283 (6.9) | 74069 (6.9) | 401171 (6.1) | 19158 (10.1) |
| Oli | 82719 (8.0) | 85342 (8.0) | 473174 (7.2) | 20452 (10.8) |
| Opc | 108157 (10.4) | 111476 (10.4) | 609520 (9.3) | 26966 (14.2) |
| All[a] | 1039252 (100.0) | 1067306 (100.0) | 6569519 (100.0) | 190005 (100.0) |

**Supplementary Table 1. Number of SNPs included in each cell-type-specific ADPRS.** The number and proportion of the post-LD shrinkage SNPs (i.e., PRS-CS-processed SNPs) included in each cell-type-specific ADPRS are shown for ROSMAP (N_SNP_ROSMAP) and A4 (N_SNP_A4). Each cell-type-specific ADPRS includes SNPs within cell-type-specific genomic regions (1,343 cell-type-specific genes per each cell type ± 30 kb margins). While the exact numbers of N_SNP_ROSMAP and N_SNP_A4 are slightly different (<5% difference due to genotype missingness in each dataset), the proportions of SNPs included in each cell-type-specific ADPRS were highly consistent. For comparison, total HRC-imputed SNP count before LD shrinkage (N_SNP_HRC) and after LD pruning (N_SNP_PRSet, p-value threshold=1) are also shown for the ROSMAP genotype data. Although LD shrinkage using PRS-CS was limited to the HapMap3 SNPs (N_SNP_ROSMAP and N_SNP_A4), it retains more SNPs with posterior effect sizes than the LD pruning approach (N_SNP_PRSet). [a]All autosomal SNPs excluding the *APOE* region.

| | Mean (s.d.) | N_nonmissing |
|---|---|---|
| AD dementia, n (%) | 538 (68.4) | 786 |
| Amyloid-β (Aβ) (sqrt) | 1.7 (1.1) | 1381 |
| Diffuse plaque (DP) (sqrt) | 0.71 (0.49) | 1452 |
| Neuritic plaque (NP) (sqrt) | 0.77 (0.53) | 1452 |
| PHFtau (sqrt) | 2.3 (1.4) | 1451 |
| Neurofibrillary tangle (NFT) (sqrt) | 0.70 (0.43) | 1452 |
| Cognitive decline | -0.017 (0.094) | 1374 |

**Supplementary Table 2. AD endophenotypes tested in ROSMAP.** The mean and standard deviation (s.d.) of the AD endophenotypes tested for their associations with cell-type-specific ADPRSs in ROSMAP are shown. For AD dementia (binary trait), we indicated the number of cases and the proportion out of the case (AD dementia) + control (cognitively unimpaired, no AD pathology) subset used for the analyses with AD dementia as the outcome (n=786). Abbreviations: N_nonmissing, number of participants with non-missing data; sqrt, square root-transformed values

| Model | OR | 95% CI | z-value | p-value | FDR |
|---|---|---|---|---|---|
| **All** | **1.53** | **1.28 to 1.85** | **4.52** | **$6.2\times10^{-6}$** | **$3.4\times10^{-5}$** |
| Ex | 1.04 | 0.87 to 1.24 | 0.44 | 0.66 | 0.68 |
| In | 1.11 | 0.92 to 1.35 | 1.12 | 0.26 | 0.35 |
| Ast | 1.18 | 0.98 to 1.42 | 1.74 | 0.082 | 0.13 |
| **Mic** | **1.45** | **1.20 to 1.75** | **3.87** | **$1.1\times10^{-4}$** | **$3.9\times10^{-4}$** |
| Oli | 1.26 | 1.05 to 1.51 | 2.47 | 0.014 | 0.030 |
| Opc | 1.05 | 0.88 to 1.25 | 0.54 | 0.59 | 0.64 |
| *APOE* ε4 | 6.81 | 4.31 to 11.2 | 7.88 | $3.3\times10^{-15}$ | NA |
| *APOE* ε2 | 0.38 | 0.24 to 0.60 | -4.17 | $3.1\times10^{-5}$ | NA |

**Supplementary Table 3. Association between cell-type-specific ADPRS and AD dementia in ROSMAP (case: n=538, control: n=248).** OR (odds ratio) of AD dementia per 1 s.d. increase in ADPRS is shown. ADPRS models were adjusted for *APOE* ε4, *APOE* ε2, age at death, sex, years of education, genotyping platform, and the first three genotype principal components. For comparison of effect sizes, ORs for *APOE* ε4 and ε2 from the same model as All-ADPRS (with the same covariates) were shown in the bottom two lines of the table. False discovery rate (FDR) correction was applied across all main tests in ROSMAP (**Supplementary Tables 3-9**), and statistically significant results (FDR<0.025) were indicated in bold. (Also see **Fig. 2**). Abbreviations: NA, not applicable.

| Model | Beta | 95% CI | t-value | p-value | FDR |
|---|---|---|---|---|---|
| **All** | **0.081** | **0.027 to 0.14** | **2.92** | **$3.6\times10^{-3}$** | **$9.3\times10^{-3}$** |
| Ex | 0.020 | -0.034 to 0.075 | 0.73 | 0.46 | 0.53 |
| In | -0.028 | -0.082 to 0.027 | -0.9835 | 0.3255 | 0.41 |
| **Ast** | **0.093** | **0.039 to 0.15** | **3.37** | **$7.8\times10^{-4}$** | **$2.6\times10^{-3}$** |
| Mic | 0.057 | $2.4\times10^{-3}$ to 0.11 | 2.05 | 0.041 | 0.074 |
| Oli | 0.055 | $1.1\times10^{-3}$ to 0.11 | 2.00 | 0.045 | 0.079 |
| Opc | 0.014 | -0.041 to 0.068 | 0.49 | 0.62 | 0.66 |
| *APOE* ε4 | 0.65 | 0.53 to 0.76 | 11.1 | $<2.0\times10^{-16}$ | NA |
| *APOE* ε2 | -0.35 | -0.50 to -0.21 | -4.73 | $2.5\times10^{-6}$ | NA |

**Supplementary Table 4. Association between cell-type-specific ADPRS and Aβ in ROSMAP (n=1,381).** Beta (effect size) corresponds to units changed in Aβ per 1 s.d. increase in ADPRS. ADPRS models were adjusted for *APOE* ε4, *APOE* ε2, age at death, sex, genotyping platform, and the first three genotype principal components. For comparison of effect sizes, the beta for *APOE* ε4 and ε2 from the same model as All-ADPRS (with the same covariates) were shown in the bottom two lines of the table. False discovery rate (FDR) correction was applied across all main tests in ROSMAP (**Supplementary Tables 3-9**), and statistically significant results (FDR<0.025) were indicated in bold. (Also see **Fig. 2**).

| Model | Beta | 95% CI | t-value | p-value | FDR |
|---|---|---|---|---|---|
| All | 0.013 | -0.011 to 0.037 | 1.04 | 0.30 | 0.39 |
| Ex | $-6.8\times10^{-3}$ | -0.031 to 0.018 | -0.55 | 0.58 | 0.64 |
| In | -0.011 | -0.036 to 0.013 | -0.92 | 0.36 | 0.43 |
| **Ast** | **0.034** | **$9.8\times10^{-3}$ to 0.058** | **2.75** | **$6.0\times10^{-3}$** | **0.014** |
| Mic | 0.016 | $-8.2\times10^{-3}$ to 0.040 | 1.30 | 0.19 | 0.26 |
| Oli | 0.026 | $1.9\times10^{-3}$ to 0.050 | 2.12 | 0.034 | 0.070 |
| Opc | 0.012 | $-1.3\times10^{-3}$ to 0.036 | 0.93 | 0.35 | 0.43 |
| *APOE* ε4 | 0.26 | 0.21 to 0.31 | 9.93 | $<2\times10^{-16}$ | NA |
| *APOE* ε2 | -0.14 | -0.21 to -0.076 | -4.28 | $2.0\times10^{-5}$ | NA |

**Supplementary Table 5. Association between cell-type-specific ADPRS and diffuse plaque burden in ROSMAP (n=1,452).** Beta (effect size) corresponds to units changed in diffuse plaque burden per 1 s.d. increase in ADPRS. ADPRS models were adjusted for *APOE* ε4, *APOE* ε2, age at death, sex, genotyping platform, and the first three genotype principal components. For comparison of effect sizes, the beta for *APOE* ε4 and ε2 from the same model as All-ADPRS (with the same covariates) were shown in the bottom two lines of the table. False discovery rate (FDR) correction was applied across all main tests in ROSMAP (**Supplementary Tables 3-9**), and statistically significant results (FDR<0.025) were indicated in bold. (Also see **Fig. 2**).

| Model | Beta | 95% CI | t-value | p-value | FDR |
|---|---|---|---|---|---|
| **All** | **0.059** | **0.036 to 0.085** | **4.53** | **$6.3\times10^{-6}$** | **$3.4\times10^{-5}$** |
| Ex | 0.020 | $-6.1\times10^{-3}$ to 0.045 | 1.50 | 0.13 | 0.19 |
| In | $1.2\times10^{-4}$ | -0.026 to 0.026 | $8.8\times10^{-3}$ | 0.99 | 0.99 |
| **Ast** | **0.051** | **0.026 to 0.077** | **3.94** | **$8.4\times10^{-5}$** | **$3.2\times10^{-4}$** |
| **Mic** | **0.055** | **0.029 to 0.080** | **4.20** | **$2.8\times10^{-5}$** | **$1.3\times10^{-4}$** |
| **Oli** | **0.056** | **0.031 to 0.082** | **4.33** | **$1.6\times10^{-5}$** | **$7.7\times10^{-5}$** |
| Opc | 0.012 | -0.014 to 0.038 | 0.90 | 0.37 | 0.43 |
| *APOE* ε4 | 0.32 | 0.27 to 0.38 | 11.8 | $<2.0\times10^{-16}$ | NA |
| *APOE* ε2 | -0.19 | -0.25 to -0.12 | -5.39 | $8.4\times10^{-8}$ | NA |

**Supplementary Table 6. Association between cell-type-specific ADPRS and neuritic plaque burden in ROSMAP (n=1,452).** Beta (effect size) corresponds to units changed in neuritic plaque burden per 1 s.d. increase in ADPRS. ADPRS models were adjusted for *APOE* ε4, *APOE* ε2, age at death, sex, genotyping platform, and the first three genotype principal components. For comparison of effect sizes, the beta for *APOE* ε4 and ε2 from the same model as All-ADPRS (with the same covariates) were shown in the bottom two lines of the table. False discovery rate (FDR) correction was applied across all main tests in ROSMAP (**Supplementary Tables 3-9**), and statistically significant results (FDR<0.025) were indicated in bold. (Also see **Fig. 2**).

| Model | Beta | 95% CI | t-value | p-value | FDR |
|---|---|---|---|---|---|
| **All** | **0.24** | **0.17 to 0.31** | **6.72** | **$2.6\times10^{-11}$** | **$1.3\times10^{-9}$** |
| **Ex** | **0.10** | **0.032 to 0.17** | **2.84** | **$4.5\times10^{-3}$** | **0.011** |
| In | 0.063 | $-7.3\times10^{-3}$ to 0.13 | 1.76 | 0.079 | 0.13 |
| **Ast** | **0.12** | **0.047 to 0.19** | **3.29** | **$1.0\times10^{-3}$** | **$3.0\times10^{-3}$** |
| **Mic** | **0.21** | **0.15 to 0.28** | **6.09** | **$1.4\times10^{-9}$** | **$2.3\times10^{-8}$** |
| **Oli** | **0.18** | **0.11 to 0.25** | **5.17** | **$2.7\times10^{-7}$** | **$2.2\times10^{-6}$** |
| Opc | 0.087 | 0.017 to 0.16 | 2.45 | 0.014 | 0.031 |
| *APOE* ε4 | 0.77 | 0.62 to 0.91 | 10.3 | $<2.0\times10^{-16}$ | NA |
| *APOE* ε2 | -0.31 | -0.50 to -0.13 | -3.35 | $8.3\times10^{-4}$ | NA |

**Supplementary Table 7. Association between cell-type-specific ADPRS and tau in ROSMAP (n=1,451).** Beta (effect size) corresponds to units changed in tau per 1 s.d. increase in ADPRS. ADPRS models were adjusted for *APOE* ε4, *APOE* ε2, age at death, sex, genotyping platform, and the first three genotype principal components. For comparison of effect sizes, the beta for *APOE* ε4 and ε2 from the same model as All-ADPRS (with the same covariates) were shown in the bottom two lines of the table. False discovery rate (FDR) correction was applied across all main tests in ROSMAP (**Supplementary Tables 3-9**), and statistically significant results (FDR<0.025) were indicated in bold. (Also see **Fig. 2**).

| Model | Beta | 95% CI | t-value | p-value | FDR |
|---|---|---|---|---|---|
| **All** | **0.068** | **0.048 to 0.089** | **6.53** | **$9.4\times10^{-11}$** | **$2.3\times10^{-9}$** |
| Ex | 0.017 | $-3.6\times10^{-3}$ to 0.038 | 1.63 | 0.10 | 0.16 |
| In | 0.015 | $-5.7\times10^{-3}$ to 0.036 | 1.42 | 0.16 | 0.22 |
| **Ast** | **0.035** | **0.014 to 0.056** | **3.33** | **$8.8\times10^{-4}$** | **$2.7\times10^{-3}$** |
| **Mic** | **0.055** | **0.035 to 0.076** | **5.30** | **$1.4\times10^{-7}$** | **$1.3\times10^{-6}$** |
| **Oli** | **0.049** | **0.029 to 0.070** | **4.70** | **$2.8\times10^{-6}$** | **$2.0\times10^{-5}$** |
| Opc | 0.022 | $1.1\times10^{-3}$ to 0.043 | 2.06 | 0.039 | 0.074 |
| *APOE* ε4 | 0.23 | 0.19 to 0.28 | 10.5 | $<2.0\times10^{-16}$ | NA |
| *APOE* ε2 | -0.12 | -0.17 to -0.066 | -4.34 | $1.5\times10^{-5}$ | NA |

**Supplementary Table 8. Association between cell-type-specific ADPRS and neurofibrillary tangle (NFT) burden in ROSMAP (n=1,452).** Beta (effect size) corresponds to units changed in neuritic plaque burden per 1 s.d. increase in ADPRS. ADPRS models were adjusted for *APOE* ε4, *APOE* ε2, age at death, sex, genotyping platform, and the first three genotype principal components. For comparison of effect sizes, the beta for *APOE* ε4 and ε2 from the same model as All-ADPRS (with the same covariates) were shown in the bottom two lines of the table. False discovery rate (FDR) correction was applied across all main tests in ROSMAP (**Supplementary Tables 3-9**), and statistically significant results (FDR<0.025) were indicated in bold. (Also see **Fig. 2**).

| Model | Beta | 95% CI | t-value | p-value | FDR |
|---|---|---|---|---|---|
| **All** | **-0.013** | **-0.018 to $-8.6\times10^{-3}$** | **-5.50** | **$4.5\times10^{-8}$** | **$5.5\times10^{-7}$** |
| Ex | $-4.2\times10^{-3}$ | $-9.0\times10^{-3}$ to $5.6\times10^{-4}$ | -1.73 | 0.084 | 0.13 |
| In | $1.5\times10^{-4}$ | $-4.7\times10^{-3}$ to $5.0\times10^{-3}$ | 0.061 | 0.95 | 0.97 |
| Ast | $-5.1\times10^{-3}$ | $-9.9\times10^{-3}$ to $-2.7\times10^{-4}$ | -2.07 | 0.038 | 0.074 |
| **Mic** | **$-9.8\times10^{-3}$** | **-0.015 to $-5.1\times10^{-3}$** | **-4.05** | **$5.5\times10^{-5}$** | **$2.3\times10^{-4}$** |
| **Oli** | **$-7.2\times10^{-3}$** | **-0.012 to $-2.5\times10^{-3}$** | **-2.96** | **$3.2\times10^{-3}$** | **$8.7\times10^{-3}$** |
| Opc | $-4.1\times10^{-3}$ | $-8.9\times10^{-3}$ to $7.3\times10^{-4}$ | -1.67 | 0.096 | 0.15 |
| *APOE* ε4 | -0.053 | -0.063 to -0.0043 | -10.3 | $<2\times10^{-16}$ | NA |
| *APOE* ε2 | 0.017 | $4.3\times10^{-3}$ to 0.030 | 2.63 | $8.7\times10^{-3}$ | NA |

**Supplementary Table 9. Association between cell-type-specific ADPRS and cognitive decline (CogDec) in ROSMAP (n=1,374).** Beta (effect size) corresponds to units changed in CogDec per 1 s.d. increase in ADPRS. ADPRS models were adjusted for *APOE* ε4, *APOE* ε2, genotyping platform, and the first three genotype principal components. For comparison of effect sizes, the beta for *APOE* ε4 and ε2 from the same model as All-ADPRS (with the same covariates) were shown in the bottom two lines of the table. False discovery rate (FDR) correction was applied across all main tests in ROSMAP (**Supplementary Tables 3-9**), and statistically significant results (FDR<0.025) were indicated in bold. (Also see **Fig. 2**).

| Model | Beta | 95% CI | t-value | p-value |
|---|---|---|---|---|
| Ex (adjusted for Mic) | 0.086 | 0.017 to 0.16 | 2.43 | 0.015 |
| Ast (adjusted for Mic) | 0.096 | 0.027 to 0.17 | 2.72 | $6.7\times10^{-3}$ |
| Oli (adjusted for Mic) | 0.091 | $7.4\times10^{-3}$ to 0.17 | 2.14 | 0.033 |

**Supplementary Table 10. Association between cell-type-specific ADPRS and tau in ROSMAP (n=1,451), adjusting for Mic-ADPRS.** Beta (effect size) corresponds to units changed in tau per 1 s.d. increase in ADPRS. ADPRS models were adjusted for Mic-ADPRS, *APOE* ε4, *APOE* ε2, age at death, sex, genotyping platform, and the first three genotype principal components.

| Model | Beta | 95% CI | t-value | p-value |
|---|---|---|---|---|
| Ex (excluding Mic) | 0.10 | 0.034 to 0.17 | 2.92 | $3.6 \times 10^{-3}$ |
| Ast (excluding Mic) | 0.092 | 0.022 to 0.16 | 2.58 | $9.9 \times 10^{-3}$ |
| Oli (excluding Mic) | 0.16 | 0.086 to 0.22 | 4.37 | $1.3 \times 10^{-5}$ |

**Supplementary Table 11. Association between cell-type-specific ADPRS and tau in ROSMAP (n=1,451), excluding genes overlapping with Mic-ADPRS.** Beta (effect size) corresponds to units changed in tau per 1 s.d. increase in ADPRS. Ex-, Ast-, and Oli- ADPRS were calculated after excluding genes overlapping with Mic-ADPRS. ADPRS models were adjusted for *APOE* ε4, *APOE* ε2, age at death, sex, genotyping platform, and the first three genotype principal components.

| | ROSMAP (n=201) |
|---|---|
| Mean Age at Death, years (SD) | 89.7 (5.5) |
| Female (%) | 126 (63) |
| Mean Education, years (SD) | 14.6 (2.6) |
| *APOE* ε4 carrier (%) | 40 (20) |
| Elevated Aβ (%) | 127 (63) |
| Pathological diagnosis of AD | 122 (61) |
| Median MMSE (IQR) | 25 (8.8) |
| All-cause dementia (%) | 76 (38) |
| AD dementia (%) | 62 (31) |
| Proportion of Activated Microglia (PAM) | 0.084 (0.057) |

**Supplementary Table 12. Study Participant Characteristics (MAP study microglial morphology subset).**

| Phenotype | Cell Type (Genomic Margin) | Beta or OR | 95% CI | t-value | p-value |
|---|---|---|---|---|---|
| AD dem | Mic (10 kb) | 1.53 | 1.27 to 1.85 | 4.37 | $1.2\times10^{-5}$ |
| AD dem | Mic (100 kb) | 1.57 | 1.30 to 1.91 | 4.67 | $3.0\times10^{-6}$ |
| Aβ | Ast (10 kb) | 0.080 | 0.026 to 0.13 | 2.92 | $3.5\times10^{-3}$ |
| Aβ | Ast (100 kb) | 0.093 | 0.039 to 0.15 | 3.37 | $7.8\times10^{-4}$ |
| DP | Ast (10 kb) | 0.032 | $7.6\times10^{-3}$ to 0.056 | 2.58 | 0.010 |
| DP | Ast (100 kb) | 0.040 | 0.016 to 0.064 | 3.25 | $1.2\times10^{-3}$ |
| NP | Ast (10 kb) | 0.042 | 0.017 to 0.068 | 3.25 | $1.2\times10^{-3}$ |
| NP | Ast (100 kb) | 0.056 | 0.031 to 0.082 | 4.33 | $1.6\times10^{-5}$ |
| NP | Mic (10 kb) | 0.051 | 0.025 to 0.076 | 3.91 | $9.8\times10^{-5}$ |
| NP | Mic (100 kb) | 0.072 | 0.046 to 0.097 | 5.56 | $3.2\times10^{-8}$ |
| NP | Oli (10 kb) | 0.060 | 0.035 to 0.086 | 4.64 | $3.7\times10^{-6}$ |
| NP | Oli (100 kb) | 0.059 | 0.034 to 0.085 | 4.58 | $5.1\times10^{-6}$ |
| Tau | Ex (10 kb) | 0.066 | $-3.8\times10^{-3}$ to 0.14 | 1.85 | 0.064 |
| Tau | Ex (100 kb) | 0.15 | 0.082 to 0.22 | 4.28 | $2.0\times10^{-5}$ |
| Tau | Ast (10 kb) | 0.11 | 0.043 to 0.18 | 3.16 | $1.6\times10^{-3}$ |
| Tau | Ast (100 kb) | 0.12 | 0.047 to 0.19 | 3.29 | $1.0\times10^{-3}$ |
| Tau | Mic (10 kb) | 0.21 | 0.14 to 0.27 | 5.85 | $6.0\times10^{-9}$ |
| Tau | Mic (100 kb) | 0.24 | 0.17 to 0.31 | 6.92 | $6.8\times10^{-12}$ |
| Tau | Oli (10 kb) | 0.18 | 0.11 to 0.25 | 5.01 | $6.0\times10^{-7}$ |
| Tau | Oli (100 kb) | 0.19 | 0.12 to 0.26 | 5.48 | $5.2\times10^{-8}$ |
| NFT | Ast (10 kb) | 0.032 | 0.011 to 0.052 | 3.02 | $2.5\times10^{-3}$ |

| | | | | | |
|---|---|---|---|---|---|
| NFT | Ast (100 kb) | 0.034 | 0.014 to 0.055 | 3.28 | $1.1\times10^{-3}$ |
| NFT | Mic (10 kb) | 0.050 | 0.029 to 0.070 | 4.78 | $2.0\times10^{-6}$ |
| NFT | Mic (100 kb) | 0.066 | 0.046 to 0.087 | 6.39 | $2.1\times10^{-10}$ |
| NFT | Oli (10 kb) | 0.048 | 0.027 to 0.068 | 4.54 | $6.1\times10^{-6}$ |
| NFT | Oli (100 kb) | 0.051 | 0.031 to 0.072 | 4.97 | $7.5\times10^{-7}$ |
| CogDec | Mic (10 kb) | -0.011 | -0.016 to $-6.7\times10^{-3}$ | -4.72 | $2.6\times10^{-6}$ |
| CogDec | Mic (100 kb) | -0.013 | -0.017 to $-7.8\times10^{-3}$ | -5.19 | $2.4\times10^{-7}$ |

**Supplementary Table 13. Association between cell-type-specific ADPRS using different genomic margins and AD endophenotypes in ROSMAP.** For the significant findings using cell-type-specific ADPRS using ± 30kb margins (FDR<0.025 in **Fig. 2**), we performed sensitivity analyses using cell-type-specific ADPRS using different genomic margins (genes ± 10kb or ± 100kb). All associations were similar to the results from ±30 kb (within 95% CI of the results reported in supplementary tables 3-9). Abbreviations: AD dem, AD with dementia; CogDec, cognitive decline; OR, odds ratio.

| Model | Effect type | Effect (95% bootstrap CI) | p-value |
|---|---|---|---|
| Ast → DP →NP | ACME | 0.023 ($6.9\times10^{-3}$ to 0.04) | $5.4\times10^{-3}$ |
| | ADE | 0.028 ($8.8\times10^{-3}$ to 0.05) | $4.0\times10^{-3}$ |
| | Total effect | 0.051 (0.026 to 0.08) | $2.0\times10^{-4}$ |
| | Mediated proportion | 0.46 (0.18 to 0.74) | $5.6\times10^{-3}$ |
| Ast → NP → NFT | ACME | 0.023 (0.012 to 0.03) | $<1.0\times10^{-4}$ |
| | ADE | 0.012 ($-4.3\times10^{-3}$ to 0.03) | 0.15 |
| | Total effect | 0.035 (0.015 to 0.05) | $2.0\times10^{-4}$ |
| | Mediated proportion | 0.65 (0.38 to 1.24) | $2.0\times10^{-4}$ |
| Mic → NP → NFT | ACME | 0.024 (0.013 to 0.03) | $<1.0\times10^{-4}$ |
| | ADE | 0.031 (0.014 to 0.05) | $<1.0\times10^{-4}$ |
| | Total effect | 0.054 (0.034 to 0.07) | $<1.0\times10^{-4}$ |
| | Mediated proportion | 0.44 (0.27 to 0.66) | $<1.0\times10^{-4}$ |
| Mic → NFT → CogDec | ACME | $-1.8\times10^{-3}$ ($-3.0\times10^{-3}$ to 0) | $5.8\times10^{-3}$ |
| | ADE | $-5.5\times10^{-3}$ ($-9.7\times10^{-3}$ to 0) | $6.0\times10^{-3}$ |
| | Total effect | $-7.3\times10^{-3}$ (-0.012 to 0) | $2.0\times10^{-4}$ |
| | Mediated proportion | 0.24 (0.080 to 0.56) | $2.0\times10^{-4}$ |

**Supplementary Table 14. Causal mediation analysis (ROSMAP).** Mediation models are run using non-parametric bootstrapping over 10,000 simulations, and 95% bootstrap confidence intervals are shown. Also see **Fig. 3**. First three models were adjusted for *APOE* ε4, ε2, age at death, sex, education, genotyping batch, and first three genotype principal components (PC1-3). The Mic → NFT → CogDec model was adjusted for neuritic plaque (NP) burden, *APOE* ε4, ε2, genotyping batch, and PC1-3. The slope of cognitive decline (CogDec) was already adjusted for age, sex, and education. Abbreviations:

ACME, average causal mediated effects. ADE, average direct effects. CogDec, cognitive decline. DP, diffuse plaque. NFT, neurofibrillary tangle. NP, neuritic plaque.

| | Mean (s.d.) | N_nonmissing |
|---|---|---|
| Aβ (SUVR) | 1.1 (0.19) | 2,921 |
| Tau (SUVR) | 1.2 (0.11) | 302 |
| HV ($mm^3$) | $3.7\times10^3$ ($4.2\times10^2$) | 1,266 |
| PACC (unit) | 0.20 (2.5) | 2,918 |

**Supplementary Table 15. AD endophenotypes tested in A4.** The mean and standard deviation (s.d.) of the AD endophenotypes tested for their associations with cell-type-specific ADPRSs in A4 are shown. Abbreviations: N_nonmissing, number of participants with non-missing data. Abbreviations: HV, hippocampal volume; PACC, Preclinical Alzheimer Cognitive Composite; SUVR, standardized uptake value ratio.

| Model | Beta | 95% CI | t-value | p-value | FDR |
|---|---|---|---|---|---|
| **All** | **0.019** | **0.012 to 0.025** | **5.73** | **$1.1\times10^{-8}$** | **$3.2\times10^{-7}$** |
| **Ex** | **$8.6\times10^{-3}$** | **$2.2\times10^{-3}$ to 0.015** | **2.62** | **$8.9\times10^{-3}$** | **0.025** |
| In | $-6.2\times10^{-4}$ | $-7.0\times10^{-3}$ to $5.8\times10^{-3}$ | -0.19 | 0.85 | 0.90 |
| **Ast** | **$9.6\times10^{-3}$** | **$3.1\times10^{-3}$ to 0.016** | **2.92** | **$3.5\times10^{-3}$** | **0.012** |
| **Mic** | **0.017** | **0.011 to 0.024** | **5.35** | **$9.3\times10^{-8}$** | **$1.3\times10^{-6}$** |
| **Oli** | **$9.9\times10^{-3}$** | **$3.5\times10^{-3}$ to 0.016** | **3.03** | **$2.5\times10^{-3}$** | **0.010** |
| Opc | $7.8\times10^{-3}$ | $1.4\times10^{-3}$ to 0.014 | 2.37 | 0.018 | 0.041 |
| *APOE* ε4 | 0.14 | 0.13 to 0.15 | 22.9 | $<2\times10^{-16}$ | NA |
| *APOE* ε2 | -0.028 | -0.046 to $-9.7\times10^{-3}$ | -2.99 | $2.8\times10^{-3}$ | NA |

**Supplementary Table 16. Association between cell-type-specific ADPRS and Aβ in A4 (n=2,921).** Beta (effect size) corresponds to units changed in florbetapir PET SUVR (cortical composite) per 1 s.d. increase in ADPRS. ADPRS models were adjusted for *APOE* ε4, *APOE* ε2, age, sex, and the first three genotype principal components. For comparison of effect sizes, the beta for *APOE* ε4 and ε2 from the same model as All-ADPRS (with the same covariates) were shown in the bottom two lines of the table. False discovery rate (FDR) correction was applied across all main tests in A4, and statistically significant results (FDR<0.025) were indicated in bold. (Also see **Fig. 4**).

| Model | Beta | 95% CI | t-value | p-value |
|---|---|---|---|---|
| Ex (adjusted for Mic) | $6.8\times10^{-3}$ | $3.6\times10^{-4}$ to 0.013 | 2.07 | 0.038 |
| Ast (adjusted for Mic) | $8.0\times10^{-3}$ | $1.6\times10^{-3}$ to 0.014 | 2.43 | 0.015 |
| Oli (adjusted for Mic) | $1.3\times10^{-3}$ | $-6.1\times10^{-3}$ to $8.7\times10^{-3}$ | 0.35 | 0.73 |

**Supplementary Table 17. Association between cell-type-specific ADPRS and Aβ in A4 (n=2,921), adjusting for Mic-ADPRS.** Beta (effect size) corresponds to units changed in florbetapir PET SUVR (cortical composite) per 1 s.d. increase in ADPRS. ADPRS models were adjusted for Mic-ADPRS, *APOE* ε4, *APOE* ε2, age, sex, and the first three genotype principal components.

| Model | Beta | 95% CI | t-value | p-value |
|---|---|---|---|---|
| Ex (excluding Mic) | $8.5\times10^{-3}$ | $2.0\times10^{-3}$ to 0.015 | 2.58 | $9.9\times10^{-3}$ |
| Ast (excluding Mic) | $7.9\times10^{-3}$ | $1.5\times10^{-3}$ to 0.014 | 2.42 | 0.016 |
| Oli (excluding Mic) | $6.3\times10^{-3}$ | $-1.1\times10^{-4}$ to 0.013 | 1.93 | 0.054 |

**Supplementary Table 18. Association between cell-type-specific ADPRS and Aβ in A4 (n=2,921; excluding genes overlapping with Mic-ADPRS).** Beta (effect size) corresponds to units changed in Aβ per 1 s.d. increase in ADPRS. Ex-, Ast-, and Oli- ADPRS were calculated after excluding genes overlapping with Mic-ADPRS. ADPRS models were adjusted for *APOE* ε4, *APOE* ε2, age at death, sex, and the first three genotype principal components.

| | A4/LEARN Tau subset (n=302) |
|---|---|
| Mean Age, years (SD) | 71.7 (4.7) |
| Female (%) | 183 (61) |
| Mean Education, years (SD) | 16.3 (2.7) |
| *APOE* ε4 carrier (%) | 164 (54) |
| Mean Florbetapir, cortical SUVR (SD) | 1.29 (0.20) |
| Mean Flortaucipir, inferior temporal SUVR (SD) | 1.53 (0.28) |
| Elevated Aβ (%) | 263 (87) |
| Median MMSE (IQR) | 29 (2) |
| AD dementia (%) | 0 (0) |

**Supplementary Table 19. Study Participant Characteristics (A4/LEARN Tau subset).**

| Model | Beta | 95% CI | t-value | p-value | FDR |
|---|---|---|---|---|---|
| **All** | **0.021** | **$8.4\times10^{-3}$ to 0.033** | **3.28** | **$1.2\times10^{-3}$** | **$6.9\times10^{-3}$** |
| Ex | $-3.2\times10^{-3}$ | -0.016 to $9.4\times10^{-3}$ | -0.50 | 0.62 | 0.72 |
| In | $1.1\times10^{-3}$ | -0.012 to 0.014 | 0.16 | 0.87 | 0.90 |
| Ast | 0.014 | $1.1\times10^{-3}$ to 0.027 | 2.14 | 0.033 | 0.067 |
| **Mic** | **0.021** | **$8.2\times10^{-3}$ to 0.033** | **3.26** | **$1.2\times10^{-3}$** | **$6.9\times10^{-3}$** |
| Oli | $9.0\times10^{-3}$ | $-3.9\times10^{-3}$ to 0.022 | 1.37 | 0.17 | 0.24 |
| Opc | $3.9\times10^{-3}$ | $-8.7\times10^{-3}$ to 0.016 | 0.61 | 0.54 | 0.66 |
| *APOE* ε4 | 0.032 | 0.011 to 0.054 | 3.02 | $2.8\times10^{-3}$ | NA |
| *APOE* ε2 | -0.045 | -0.087 to $-3.8\times10^{-3}$ | -2.15 | 0.033 | NA |

**Supplementary Table 20. Association between cell-type-specific ADPRS and tau in A4 (n=302).** Beta (effect size) corresponds to units changed in flortaucipir PET SUVR (temporal lobe composite) per 1 s.d. increase in ADPRS. ADPRS models were adjusted for *APOE* ε4, *APOE* ε2, age, sex, and the first three genotype principal components. For comparison of effect sizes, the beta for *APOE* ε4 and ε2 from the same model as All-ADPRS (with the same covariates) were shown in the bottom two lines of the table. False discovery rate (FDR) correction was applied across all main tests in A4, and statistically significant results (FDR<0.025) were indicated in bold. (Also see **Fig. 3**).

| Model | Beta | 95% CI | t-value | p-value |
|---|---|---|---|---|
| Ex | 0.048 | -0.032 to 0.13 | 1.18 | 0.24 |
| Ast | 0.013 | -0.067 to 0.093 | 0.31 | 0.76 |
| Mic | 0.16 | 0.081 to 0.24 | 3.93 | $1.0\times10^{-4}$ |
| Oli | 0.057 | -0.027 to 0.14 | 1.33 | 0.18 |

**Supplementary Table 21. Association between cell-type-specific ADPRS and tau in ROSMAP CU subset (n=454).** Beta (effect size) corresponds to units changed in Aβ per 1 s.d. increase in ADPRS. ADPRS models were adjusted for *APOE* ε4, *APOE* ε2, age at death, sex, genotyping platform, and the first three genotype principal components.

| | A4/LEARN MRI subset (n=1266) |
|---|---|
| Mean Age, years (SD) | 71.5 (4.7) |
| Female (%) | 753 (59) |
| Mean Education, years (SD) | 16.7 (2.6) |
| *APOE* ε4 carrier (%) | 609 (48) |
| Mean Florbetapir, cortical SUVR (SD) | 1.22 (0.22) |
| Mean HV, $mm^3$ (SD) | 3774 (417) |
| Elevated Aβ (%)[a] | 849 (67) |
| Median MMSE (IQR) | 29 (2) |
| AD dementia (%) | 0 (0) |

**Supplementary Table 22. Study Participant Characteristics (A4/LEARN structural MRI subset).** Abbreviations: APOE, apolipoprotein E; HV, hippocampal volume; IQR, interquartile range; MMSE, Mini-Mental State Examination; SD, standard deviation; SUVR, standardized uptake value ratio (whole cerebellar reference). [a]n=1265 with data.

| Model | Beta | 95% CI | t-value | p-value | FDR |
|---|---|---|---|---|---|
| **All** | **-33** | **-52 to -14** | **-3.48** | **$5.3{\times}10^{-4}$** | **$4.9{\times}10^{-3}$** |
| Ex | -14 | -34 to 4.8 | -1.47 | 0.14 | 0.21 |
| In | 3.1 | -16 to 22 | 0.32 | 0.75 | 0.84 |
| Ast | -15 | -34 to 4.3 | -1.52 | 0.13 | 0.21 |
| Mic | -15 | -34 to 4.2 | -1.53 | 0.13 | 0.21 |
| Oli | -23 | -42 to -4.1 | -2.39 | 0.017 | 0.041 |
| Opc | -14 | -33 to 4.6 | -1.49 | 0.14 | 0.21 |
| *APOE* ε4 | -54 | -86 to -22 | -3.32 | $9.2{\times}10^{-4}$ | NA |
| *APOE* ε2 | -24 | -84 to 35 | -0.81 | 0.42 | NA |

**Supplementary Table 23. Association between cell-type-specific ADPRS and hippocampal volume (HV) in A4 (n=1,266).** Beta (effect size) corresponds to units changed in HV ($mm^3$) per 1 s.d. increase in ADPRS. ADPRS models were adjusted for *APOE* ε4, *APOE* ε2, age, sex, intracranial volume (ICV), and the first three genotype principal components. For comparison of effect sizes, the beta for *APOE* ε4 and ε2 from the same model as All-ADPRS (with the same covariates) were shown in the bottom two lines of the table. False discovery rate (FDR) correction was applied across all main tests in A4, and statistically significant results (FDR<0.025) were indicated in bold. (Also see **Fig. 3**).

| Model | Beta | 95% CI | t-value | p-value | FDR |
|---|---|---|---|---|---|
| **All** | **-0.13** | **-0.21 to -0.045** | **-3.02** | **$2.5\times10^{-3}$** | **0.010** |
| Ex | -0.027 | -0.11 to 0.056 | -0.64 | 0.52 | 0.66 |
| In | -0.095 | -0.18 to -0.013 | -2.26 | 0.024 | 0.051 |
| **Ast** | **-0.12** | **-0.20 to -0.037** | **-2.83** | **$4.6\times10^{-3}$** | **0.014** |
| Mic | -0.088 | -0.17 to $-5.6\times10^{-3}$ | -2.10 | 0.036 | 0.068 |
| Oli | $-2.2\times10^{-3}$ | -0.084 to 0.080 | -0.053 | 0.96 | 0.96 |
| Opc | -0.053 | -0.14 to 0.030 | -1.26 | 0.21 | 0.28 |
| *APOE* ε4 | -0.26 | -0.42 to -0.11 | -3.42 | $6.4\times10^{-4}$ | NA |
| *APOE* ε2 | -0.013 | -0.25 to 0.22 | -0.11 | 0.91 | NA |

**Supplementary Table 24. Association between cell-type-specific ADPRS and Preclinical Alzheimer Cognitive Composite (PACC) in A4 (n=2,918).** Beta (effect size) corresponds to units changed in PACC per 1 s.d. increase in ADPRS. ADPRS models were adjusted for *APOE* ε4, *APOE* ε2, age, sex, years of education, and the first three genotype principal components. For comparison of effect sizes, the beta for *APOE* ε4 and ε2 from the same model as All-ADPRS (with the same covariates) were shown in the bottom two lines of the table. False discovery rate (FDR) correction was applied across all main tests in A4, and statistically significant results (FDR<0.025) were indicated in bold. (Also see **Fig. 3**).

| Phenotype | Cell Type (Genomic Margin) | Beta or OR | 95% CI | t-value | p-value |
|---|---|---|---|---|---|
| Aβ | Ex (10 kb) | $8.5\times10^{-3}$ | $2.1\times10^{-3}$ to 0.015 | 2.61 | $9.1\times10^{-3}$ |
| Aβ | Ex (100 kb) | $8.6\times10^{-3}$ | $2.2\times10^{-3}$ to 0.015 | 2.62 | $8.9\times10^{-3}$ |
| Aβ | Ast (10 kb) | $8.0\times10^{-3}$ | $1.6\times10^{-3}$ to 0.014 | 2.45 | 0.014 |
| Aβ | Ast (100 kb) | $9.6\times10^{-3}$ | $3.1\times10^{-3}$ to 0.016 | 2.92 | $3.5\times10^{-3}$ |
| Aβ | Mic (10 kb) | 0.015 | $8.2\times10^{-3}$ to 0.021 | 4.48 | $7.8\times10^{-6}$ |
| Aβ | Mic (100 kb) | 0.017 | 0.011 to 0.024 | 5.35 | $9.3\times10^{-8}$ |
| Aβ | Oli (10 kb) | $9.0\times10^{-3}$ | $2.6\times10^{-3}$ to 0.015 | 2.75 | $5.9\times10^{-3}$ |
| Aβ | Oli (100 kb) | $9.9\times10^{-3}$ | $3.5\times10^{-3}$ to 0.016 | 3.03 | $2.5\times10^{-3}$ |
| Tau | Mic (10 kb) | 0.016 | $3.3\times10^{-3}$ to 0.28 | 2.50 | 0.013 |
| Tau | Mic (100 kb) | 0.020 | $7.8\times10^{-3}$ to 0.033 | 3.19 | $1.6\times10^{-3}$ |
| PACC | Ast (10 kb) | -0.11 | -0.20 to -0.031 | -2.69 | $7.2\times10^{-3}$ |
| PACC | Ast (100 kb) | -0.12 | -0.20 to -0.033 | -2.76 | $5.9\times10^{-3}$ |

**Supplementary Table 25. Association between cell-type-specific ADPRS using different genomic margins and AD endophenotypes in A4.** For the significant findings using cell-type-specific ADPRS using ± 30kb margins (FDR<0.025 in **Fig. 2**), we performed sensitivity analyses using cell-type-specific ADPRS using different genomic margins (genes ± 10kb or ± 100kb).
